# Associations of ADHD symptom severity, sleep/circadian factors, depression and quality of life: Secondary analyses of the Netherlands Sleep Registry study

**DOI:** 10.1101/2025.02.04.25321678

**Authors:** Siddhi Nair, Neha Deshpande, Catherine M. Hill, Samuele Cortese, Eus Van Someren, Sarah L. Chellappa

**Affiliations:** Centre for Innovation in Mental Health, School of Psychology, Faculty of Environmental and Life Sciences, University of Southampton, Southampton, United Kingdom; School of Clinical and Experimental Sciences, Faculty of Medicine, University of Southampton, Southampton, UK; Department of Sleep Medicine, Southampton Children’s Hospital, University Hospital Southampton NHS Foundation Trust, Southampton, United Kingdom; Solent NHS Trust, Southampton, United Kingdom; Hassenfeld Children’s Hospital at NYU Langone, New York University Child Study Center, New York City, New York, United States; DiMePRe-J-Department of Precision and Regenerative Medicine-Jonic Area, University of Bari "Aldo Moro", Bari, Italy; Netherlands Institute for Neuroscience, Department of Sleep and Cognition, Amsterdam, the Netherlands; Departments of Integrative Neurophysiology and Psychiatry, Center for Neurogenomics and Cognitive Research, VU University, Amsterdam UMC, Amsterdam Neuroscience, Amsterdam, the Netherlands

**Keywords:** Attention-deficit hyperactive disorder (ADHD), sleep disturbances, sleep complaints, insomnia, sleep quality, circadian preference, mental health, anxiety, depression, quality of life

## Abstract

**Study objectives:** We investigated whether sleep disruption and circadian preference mediate the associations of ADHD symptom severity with depression symptom severity and quality of life.

**Methods:** 1364 participants (mean: 51.86 [*SD* = 0.37] years, 75% females) from a large-scale cross-sectional online survey (Netherlands Sleep Registry) completed a sociodemographic questionnaire, the Adult ADHD Rating Scale, Hospital Anxiety and Depression Scale (HADS), Satisfaction With Life Scale (SLS) and Cantril Ladder (CL) (quality of life measures), Insomnia Severity Index, Pittsburgh Sleep Quality Index, and Munich Chronotype Questionnaire.

**Results:** Higher ADHD symptom severity was significantly associated with depression severity (*p* = 0.03), lower quality of life (*p* < 0.001), insomnia severity (*p* < 0.001), lower sleep quality (*p* < 0.001), and later circadian preference (*p* = 0.01). No sleep or circadian factor significantly mediated the association of the severity of symptoms of ADHD and depression (all *p* > 0.1). Conversely, only insomnia severity significantly mediated the association of the severity of symptoms of ADHD and quality of life (SLS: standardized beta = -0.10, 95% CI = [-0.12, -0.04]; CL: standardized beta = .103, 95% CI = [0.04, 0.16]).

**Conclusion:** ADHD symptom severity was associated with lower quality of life, primarily mediated by insomnia symptom severity. Future studies targeting insomnia complaints in this population may help mitigate their depression complaints and improve their quality of life.

## INTRODUCTION

Attention Deficit Hyperactivity Disorder (ADHD) is a common neurodevelopmental condition with a worldwide-pooled prevalence of 5-7%^1,2^. ADHD is a common neurodevelopmental condition characterized by a core of traits including challenges with attention, activity levels and impulsivity^3^. ADHD often co-occurs with mental disorders, particularly depression^4^, whereby up to 80% of adults with ADHD (18-35 years) report at least one episode of depression throughout their lifespan compared to age- and sex-matched individuals with neurotypical development^5^. A meta-analysis of 18 cross-sectional studies (1615 individuals with ADHD, 3128 individuals with neurotypical development) showed a significant association between ADHD and depression among young adults (OR: 4.66, 95% CI [2.92–7.45])^6^. Similarly, quality of life (QoL) may be affected by ADHD symptom severity^7^. A recent meta-analysis including 23 studies of children with ADHD (4-18 years) showed that both parent reported (Hedges’ *g* −1.67, 95% CI [−2.57, −0.78]) and child reported (Hedges’ *g* −1.28, 95% CI [−2.01, −0.56]) health-related QoL was lower compared to children with neurotypical development^7^. Despite the high prevalence of ADHD and its impact on public health, biological factors contributing to these associations remain to be fully established. Of note, most studies on ADHD have primarily focused on children and adolescents. Thus, it is largely unknown if middle-aged and older people with ADHD also experience a heightened likelihood for depression risk and low quality of life.

Importantly, at least 25% of adults with ADHD self-report sleep disorders, including delayed sleep-wake phase syndrome, restless legs syndrome, daytime sleepiness, and insomnia^8^. The latter is the most common sleep disorder affecting the general population, with estimates of 43-83% among ADHD^9^. A meta-analysis from 16 cross-sectional studies (722 children with ADHD and 638 age- and sex-matched children with neurotypical development) indicates higher bedtime resistance (z = 6.94), greater sleep onset difficulties (*z* = 9.38), night awakenings (*z* = 2.15), and daytime sleepiness (*z* = 1.96) in children with ADHD^10^. Similarly, a meta-analysis of 13 cross-sectional studies revealed that adults with ADHD had longer sleep onset latency (SMD = 0.80, 95% CI [0.46, 1.14]) and lowered sleep efficiency [SMD = -0.68, 95% CI [-1.03, -0.34]), compared to individuals with neurotypical development^11^. Sleep disruption/curtailment may exert multiple impacts on neurobehavioral and cognitive systems, including attention and emotional regulation^12^, which may suggest that sleep-related factors play a potential role in ADHD symptom severity. Further, sleep disruption may arise from ADHD-related impulsivity and hyperactivity; thus, sleep problems may be part of the symptomatology of ADHD. These findings collectively speak to a negative cycle involving sleep disturbances and core ADHD characteristics and symptoms. Similarly, ADHD symptom severity may be associated with circadian disturbances. Studies of circadian rhythms in adults with ADHD indicate that ∼30% report late circadian preference, delayed circadian phase (as indexed by dim light melatonin onset), and higher risk of delayed sleep-wake phase syndrome, suggesting an important role in ADHD symptom severity^13^. Sleep and circadian rhythm disturbances (SCRD) are bidirectionally intertwined with almost every category of mental disorder in a transdiagnostic manner^14^. However, the majority of these studies focus on individuals with neurotypical development. Thus, it is currently unknown whether SCRD mediates the increased risk of depression and lower quality of life in individuals with ADHD traits.

Here, we investigated whether disrupted sleep (e.g., insomnia, and low self-reported sleep quality) mediates the association of self-rated ADHD symptom severity with depression and low QoL. We utilized data from the Netherlands Sleep Registry (NSR), a large-scale online survey assessment platform with over 10,000 participants aged 18y and above. Our main hypothesis is that the associations of ADHD symptom severity with depression symptom severity and low quality of life are mediated by the severity of symptoms of insomnia, low self-reported sleep quality, and late circadian preference.

## METHODS

### Participants

Data included here are from the Netherlands Sleep Registry (NSR), an online survey dataset with > 10,000 participants (aged 18 and older) from the general population (www.slaapregister.nl). Participants were recruited via media, advertisements, and flyers distributed at healthcare institutions and conventions. The participation of people was voluntary, and all data were recorded anonymously^15^. The only inclusion criterion was an age of 18 years or older. Participants completed a variable number of randomly selected questionnaires (of 34 online questionnaires) at their convenience, resulting in a varying number and set of completed questionnaires between participants. Volunteers of the NSR participated anonymously without revealing their names and addresses, and were not exposed to interventions or behavioral constraints. The NSR therefore does not fall under the Dutch Medical Research Involving Human Subjects Act, and signed informed consent is not mandatory, as confirmed by the Medical Ethical Committee of the Academic Medical Center of the University of Amsterdam as well as the Central Committee on Research Involving Human Subjects (CCMO), The Hague, The Netherlands. Before performing this secondary data analysis, we obtained ethical approval by the University of Southampton Research Ethics Committee.

In the present study, of the 3571 that completed ADHD trait questionnaire, 1364 also fully completed the questionnaire battery of interest, which included questionnaires about insomnia (*N* = 2354), sleep quality (*N* = 2348), circadian preference (*N* = 1913), depression (*N* = 2490) and QoL (*N* = 3251) (**Table 1**).

**Table 1.** Sociodemographic and Study Characteristics (N = 1364).

| Demographic variables | M (95% CI) |
| --- | --- |
| Sex | 0.8 [0.74, 0.78] |
| Age | 51.9 [51.13, 52.59] |
| Marital status | 2.2 [2.09, 2.22] |
| Educational level | 6.6 [6.50, 6.65] |
| Employment | 3.7 [3.55, 3.83] |
| Adult ADHD Self-Report Scale | 1.7 [1.59, 1.75] |
| Hospital Anxiety and Depression Scale | 9.1 [9.00, 9.17] |
| Satisfaction with Life Scale | 23.6 [23.25, 24.0] |
| Insomnia Severity Index | 9.4 [8.99, 9.74] |
| Pittsburg Sleep Quality Index | 7.2 [6.96, 7.44] |
| Munich Chronotype Questionnaire | 3.9 [3.80, 3.92] |

### Measurements

#### ADHD

ADHD symptom severity was measured using the Adult ADHD Self-Report Scale^16^. This tool was designed to help screen adults (aged 18 years and older) with and without ADHD traits, and is validated with the full version of the ASRS (an 18-item measure corresponding to the ADHD symptoms in the DSM-IV)^16^. The current measure consists of 6-items (four on inattention and two on hyperactivity) and participants were required to rate the severity of each symptom on a five-point Likert scale (0 = never; 1 = rarely; 2 = sometimes; 3 = often; 4 = very often). The participants were classified based on their scores into no symptom group (ASRS symptom score = 0), moderate symptom group (ASRS symptom score = 1–3), and severe symptom group (ASRS symptom score = ≥ 4). The ASRS has moderate sensitivity (68.7%), high specificity (99.5%), and high total classification accuracy (97.9%). The cut-off score for potential ADHD was 4 and above.

#### Depression

Depression symptom severity was assessed using the Hospital Anxiety and Depression Scale (HADS). This 14-item self-reported scale includes 7 items for anxiety and 7 for depression with responses scored on a 4-point Likert scale, from 0-3, with higher scores representing worse symptoms. The maximum score attainable for each sub-scale is 21, with the cutoff score for the presence of the condition at 8^17^. A score of 0-7 is in the normal range, 8-10 is mild-to-moderate and 11≥ is severe.

#### QoL measures

The Satisfaction with Life scale (SLS) measures psychological well-being, and is a short 5-item questionnaire aiming to measure cognitive judgements of an individual’s life satisfaction and does not measure affect (either positive or negative)^18^. The initial 3 items focus on satisfaction with life currently and positive evaluations, whereas items 4 and 5 investigate past life satisfaction. Each item is scored on a 7-point scale ranging from 1 (strongly disagree) to 7 (strongly agree). The maximum score attainable is 35 with cutoff scores as follows: 5-9 (extremely dissatisfied), 10-14 (dissatisfied), 15-19 (slightly dissatisfied), 20 (neutral), 21-25 (slightly satisfied), 26-30 (satisfied) and 31-35 (extremely satisfied).

The Cantril’s Self-Anchoring Ladder of Life satisfaction, more commonly known as the Cantril Ladder (CL), is used to measure life satisfaction among adults. Participants were asked to visualize their life in the best possible manner and explain their dreams and hopes for the future. This is followed by asking them the opposite, that is explaining and describing their worst fears and envisioning the worst-case scenarios, and the negative emotions that come with it^19^. After the initial assessment, the individual was presented with the picture of a ladder numbered from zero to ten representing worst and best life satisfaction, respectively. The participant was asked to think about their life presently and place themselves on the ladder, indicating their current standpoint. Further, they are also asked to rate themselves from 5 years ago and where they would see themselves 5 years from now. A score of four or below indicates ‘suffering’ and scoring more than seven equates to ‘thriving’. The higher the score, the better the individual’s life satisfaction and well-being.

#### Insomnia

The Insomnia Severity Index (ISI) is a 7-item questionnaire where participants must choose options on a 5-point Likert scale ranging from 0 (not at all) to 4 (very severe/very much). The total score ranges between 0-28 with the cutoff score for likely insomnia being at least 10, and above 15 suggesting high risk of clinical insomnia indicating severe sleep disturbance and associated daytime symptoms^20^.

#### Sleep quality

The Pittsburg Sleep Quality Index (PSQI) assessed difficulties initiating and maintaining sleep^21^. This 19-item self-report questionnaire includes 7 sub-scales: sleep latency, sleep duration, subjective sleep quality, habitual sleep efficiency, sleep disturbances, daytime dysfunction, and use of sleeping medication. The questionnaire comprises both Likert type and open-ended questions, and participants must answer how often over the past month they experienced sleep difficulties and rate their overall sleep quality. Scores for each question range from 0 (no difficulty) to 3 (severe), with the cutoff score of 5, and the values above the cutoff to indicate severe sleep difficulties. The maximum score possible on the questionnaire is 21.

#### Circadian preference

The Munich Chronotype Questionnaire (MCTQ) is a self-rated scale that assesses an individual’s entrainment of working and work-free days It consists of 17 items, divided into 4 categories (work schedule, workday sleep schedule, free day sleep schedule, self-assessment of circadian preference), regarding sleep-wake behavior of individuals, attempting to distinguish weekday and weekend schedules. The MCTQ was designed to assess circadian preference by evaluating the corrected midsleep on free days, which is the midpoint between sleep onset and offset on free days^22^.

### Data analysis

Data was analyzed using IBM Statistics v.29 and SAS 9.4. Outliers (i.e., values over three standard deviations of the mean) were removed from the analyses. To standardize the dataset, all of the data were *z*-transformed. The data did not present with multicollinearity and neither did it have independent errors. Firstly, means and standard deviations were calculated for the sociodemographic data (sex, age, marital status, education level, and employment status) and the data obtained from the ASRS, ISI, PQSI, MCTQ, HADS, SLS and CL questionnaires. Additionally, percentages were also calculated for each category of the sociodemographic data. Next, we used Spearman correlations to assess the associations of ADHD symptom severity, depression symptom severity, QoL, insomnia severity, self-reported sleep quality and circadian preference. Linear and hierarchical regression models were performed to determine the effect of ADHD symptom severity on depression symptom severity and QoL, while accounting for sociodemographic factors, insomnia severity, self-reported sleep quality, and circadian preference. Hierarchical regression models were used to test the predictor significance of each variable. The sociodemographic covariates sex, age, marital status, education level and employment status were entered into the first model, followed by the addition of the sleep/circadian and ADHD symptom severity into the second model. The effect sizes for each were determined by the R^2^ values with 0.02, 0.13, and 0.26 indicating small, medium and large effect sizes, respectively. Finally, a mediation analysis was done using PROCESS (version 4, model 6: Hayes 2022) to identify whether sleep/circadian factors (i.e., self-reported sleep quality, insomnia severity, and circadian preference) mediated depression symptom severity and quality of life among individuals with ADHD traits. Two mediation analysis models were constructed: the first model included the sleep/circadian predictors self-reported sleep quality, circadian preference, and insomnia severity. In the next step, we added the covariates sex, age, marital status, education level, and employment status to create the second model. Of note, the addition of the covariates was justified by a significant increase in model fit relative to the loss of parsimony (using the Akaike information criterion, AIC).

## RESULTS

### Correlational analysis

Spearman’s rho bivariate correlations indicated a significant albeit weak association between depression symptom severity and ADHD symptom severity (*r* = 0.11, *p* = 0.03), and a robust association of QoL and ADHD symptom severity (Satisfaction with Life scale: *r* = -0.24, *p* < 0.001; Cantril Ladder: *r* = -0.24, *p* < 0.001). Thus, higher ADHD symptom severity was associated with higher depression symptom severity and lower QoL (**Figure 1**). Likewise, we observed a robust association of ADHD symptom severity with insomnia severity (*r* = 0.27, *p* < 0.001), self-reported sleep quality (*r* = 0.24, *p* < 0.001), and a significant, albeit weak, association with circadian preference (*r* = 0.08, *p* = 0.01), indicating that higher ADHD symptom severity were associated with higher insomnia severity, lower sleep quality and later circadian preference (**Figure 2**).

**Figure 1.**
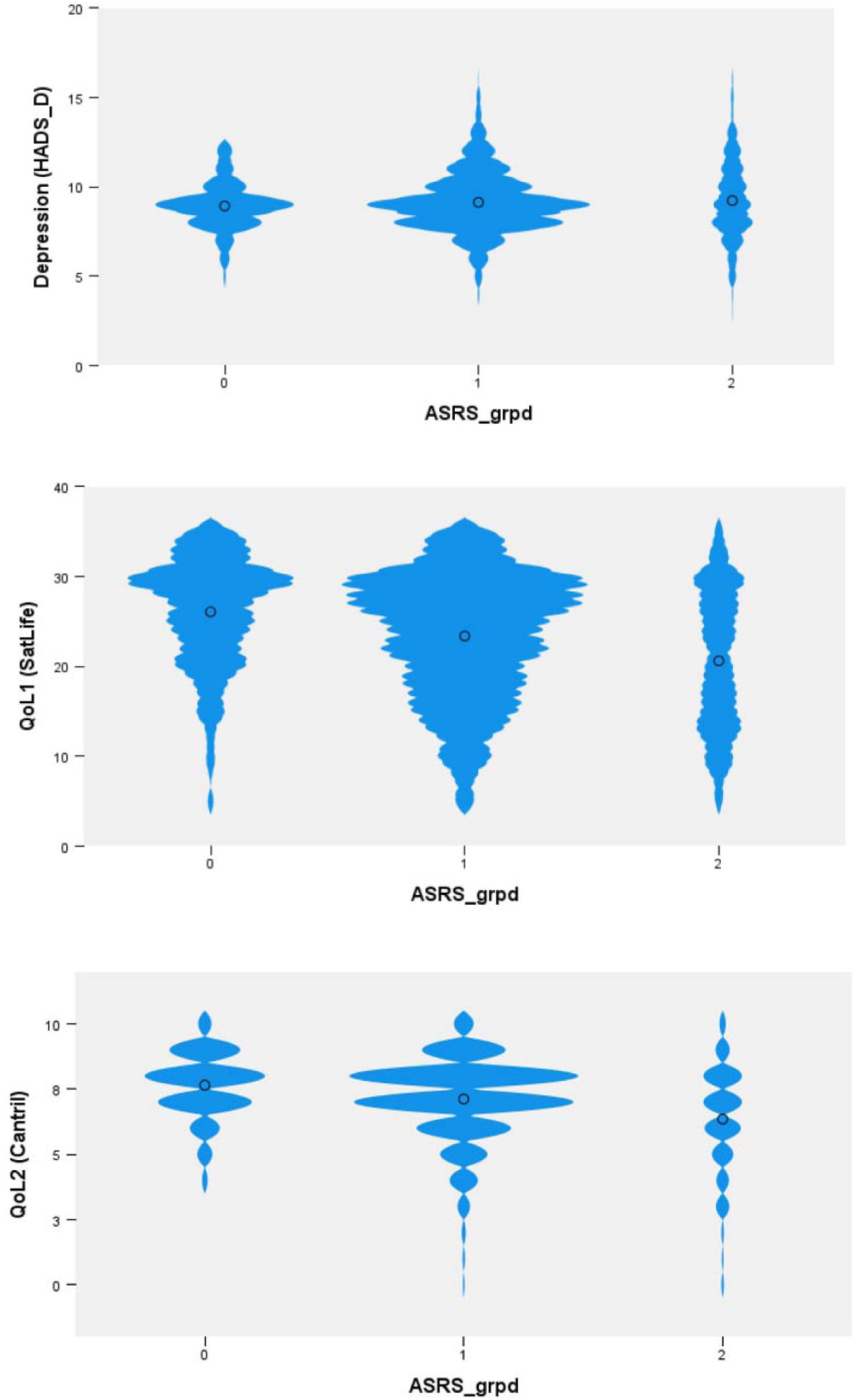
Violin plots of the associations of ADHD symptom severity with depression severity and quality of life. Higher ADHD symptom severity was associated with higher depression symptom severity (Hospital Anxiety and Depression Scale, upper panel) and with lower quality of life (Satisfaction with Life scale, middle panel; Cantril Ladder, bottom panel). The x-axis indicates the Adult ADHD Self-Report Scale grouped based on symptom severity according to the questionnaire’s cutoff thresholds (0= no symptoms, 1= moderate symptoms, 2= severe symptoms), and the y-axis indicates the corresponding depression and quality of life questionnaire scores. Note: for the upper panel, higher levels indicate higher depression symptom severity, and for the middle and bottom panels, higher levels indicate better quality of life.

**Figure 2.**
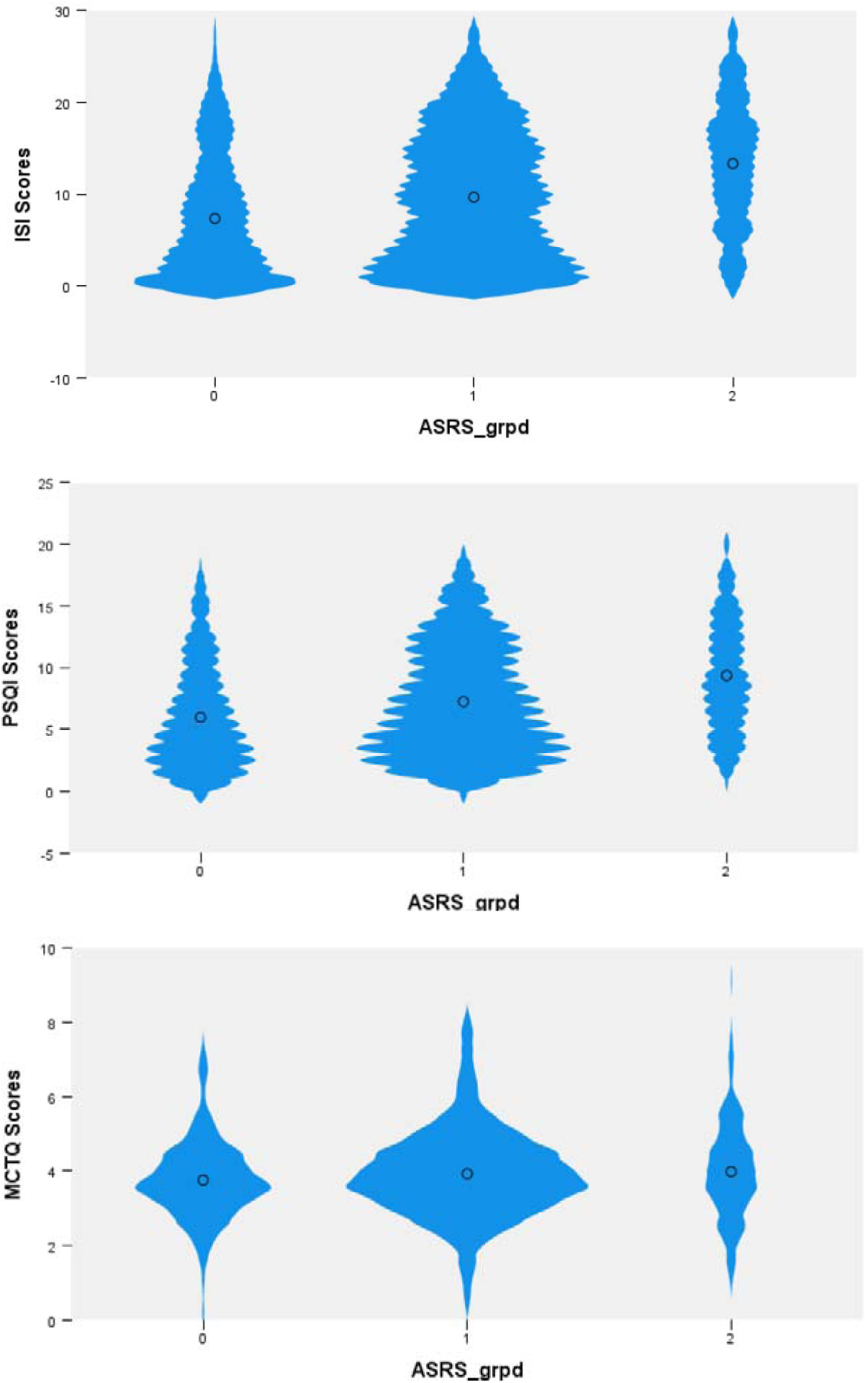
Violin plots of the associations of ADHD symptom severity with sleep and circadian factors. Higher ADHD symptom severity was associated with higher insomnia severity (Insomnia Severity Index, upper panel), lower self-reported sleep quality (Pittsburg Sleep Quality Index, middle panel), and later circadian preference (Munich Chronotype Questionnaire, bottom panel). The x-axis indicates the Adult ADHD Self-Report Scale grouped based on symptom severity according to the questionnaire’s cutoff thresholds (0= no symptoms, 1= moderate symptoms, 2= severe symptoms), and the y-axis indicates the corresponding sleep and circadian questionnaires. Note: for the upper panel, higher levels indicate higher insomnia severity, for the middle panel, higher levels indicate worse self-reported sleep quality, and for the bottom panel, higher levels indicate later circadian preference.

### Hierarchical regression models

Hierarchical multiple linear regression was carried out to identify the effects of ADHD symptom severity on depression symptom severity and QoL, controlling for covariates of interest (model 1: sociodemographic variables age, sex, marital status, educational level, employment; model 2: sociodemographic variables and sleep/circadian predictors self-reported sleep quality, circadian preference and insomnia severity).

#### Depression

None of the sociodemographic predictors significantly contributed to depression severity (model 1: *R^2^* = 0.07; all *p>*0.1; model 2: *R^2^* = 0.07; all *p>*0.1). Likewise, none of the sleep/circadian predictors (model 2: *R^2^* = 0.07; all *p>*0.09) nor ADHD symptom severity (model 2: *R^2^* = 0.07; *p*=0.11) significantly contributed to depression severity (**Table 2**).

**Table 2.**
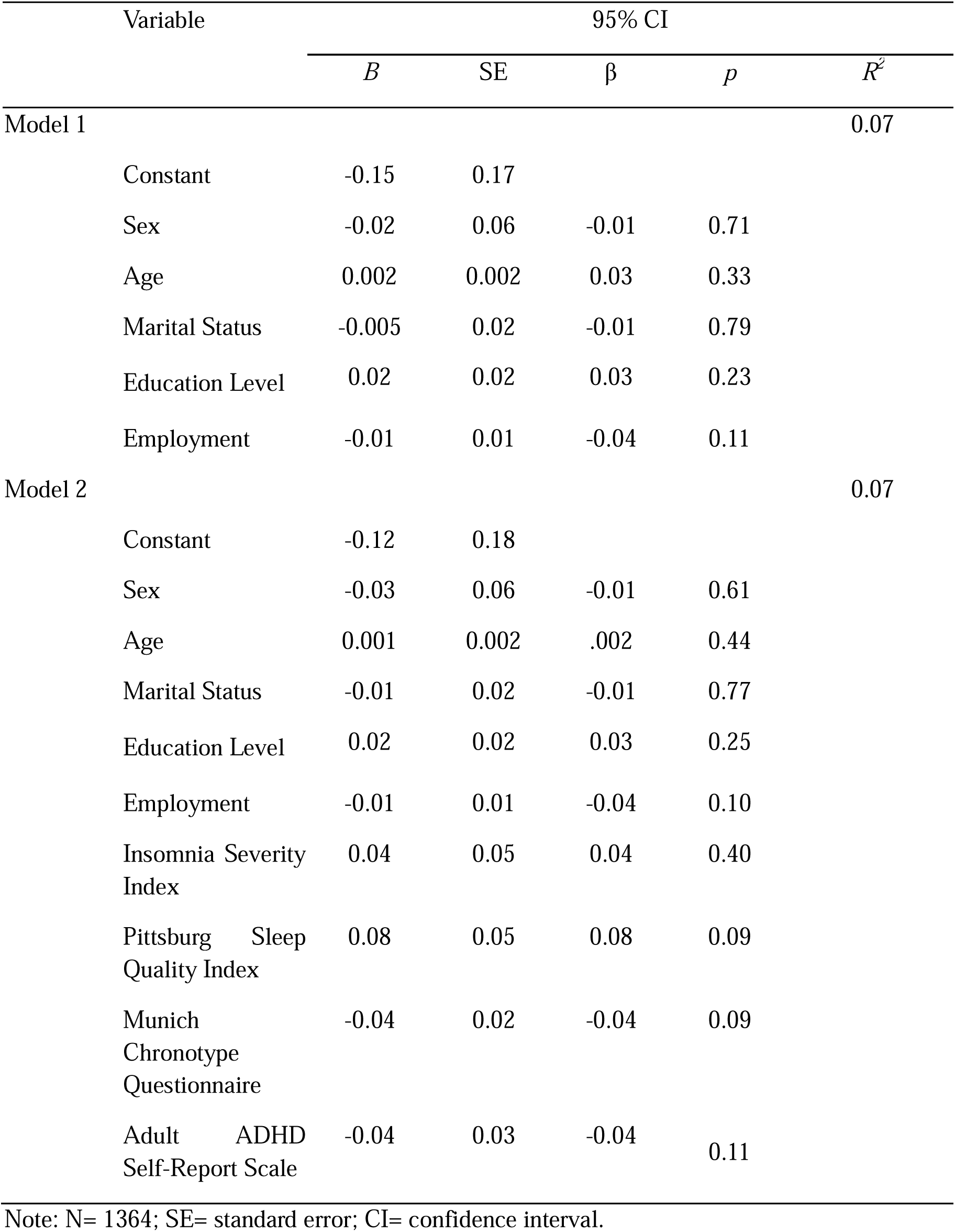
Hierarchical Regression Results for depression symptom severity.

#### Quality of life

##### Satisfaction with Life Scale (SLS)

None of the sociodemographic predictors significantly contributed to SLS-related quality of life (model 2: *R^2^* = 0.35; *p >* 0.09). Conversely, insomnia severity and ADHD symptom severity were significant contributors of SLS-related quality of life (model 2: *R^2^* = 0.35; respectively, *p* < 0.001 and *p=* 0.03). Overall, when only sociodemographic covariates were included (model 1) these variables explained 26% of the variance (medium effect size). In contrast, a model including sociodemographic, sleep/circadian predictors and ADHD symptom severity (model 2) accounted for 35% of the variance (large effect size) (**Table 3**).

**Table 3.**
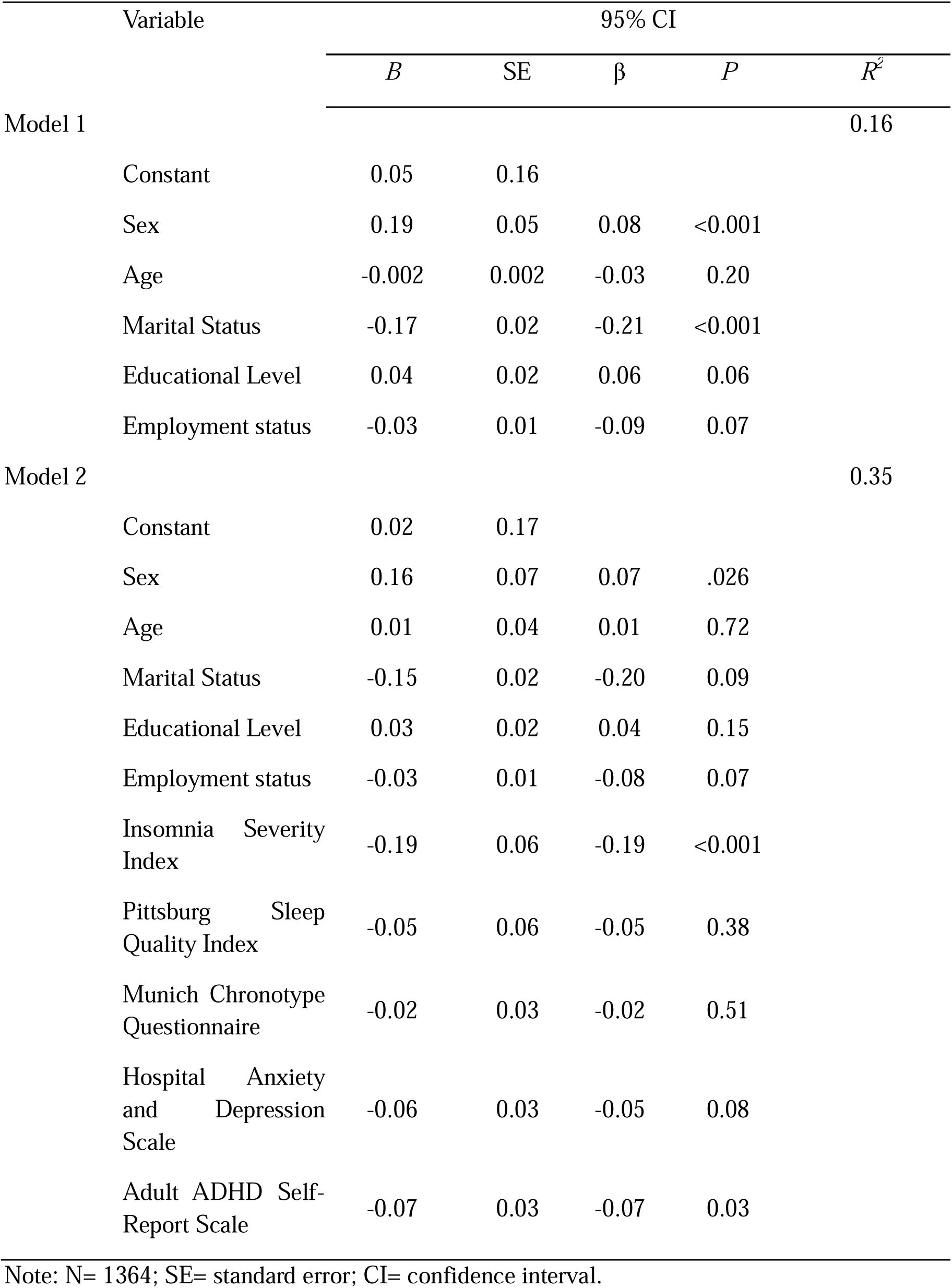
Hierarchical regression results for the Satisfaction with Life scale.

##### Cantril Ladder questionnaire (CL)

The sociodemographic predictors sex, age, marital status and educational level significantly contributed to CL-related quality of life (model 2: *R^2^* = 0.27; *p <* 0.01). Moreover, insomnia severity, depression symptom severity, and ADHD symptom severity significantly contributed to CL-related quality of life (respectively, *p* = 0.002, *p* < 0.03, and *p=* 0.03). Overall, when only sociodemographic covariates were included (model 1) these variables explained 16% of the variance (medium effect size). In contrast, a model including sociodemographic, sleep/circadian predictors and ADHD symptom severity (model 2) accounted for 27% of the variance (large effect size) (**Table 4**).

**Table 4.** Hierarchical regression results for the Cantril Ladder questionnaire.

| Variable | 95% CI | | | | $R^2$ |
| --- | --- | --- | --- | --- | --- |
| | *B* | *SE* | $\beta$ | *P* | |
| Model 1 |  |  |  |  | 0.16 |
| Constant | 1.45 | .08 |  |  |  |
| Sex | -.08 | .03 | -0.08 | 0.02 |  |
| Age | -.05 | .02 | -0.10 | 0.004 |  |
| Marital Status | .03 | .01 | 0.09 | 0.004 |  |
| Education Level | -.02 | .01 | -0.08 | 0.015 |  |
| Employment status | .00 | .01 | 0.03 | 0.41 |  |
| Model 2 |  |  |  |  | 0.27 |
| Constant | 1.45 | .08 |  |  |  |
| Sex | -.07 | .03 | -0.07 | 0.03 |  |
| Age | -.04 | .02 | -0.09 | 0.009 |  |
| Marital Status | .03 | .01 | 0.09 | 0.003 |  |
| Education Level | -.02 | .01 | -0.08 | 0.01 |  |
| Employment | .00 | .01 | 0.03 | 0.39 |  |
| Insomnia Severity Index | .08 | .03 | 0.19 | 0.002 |  |
| Pittsburg Sleep Quality Index | .01 | .03 | 0.02 | 0.71 |  |
| Munich Chronotype Questionnaire | .01 | .01 | 0.01 | 0.67 |  |
| Hospital Anxiety and Depression Scale | .03 | .01 | 0.07 | 0.03 |  |
| Adult ADHD Self-Report Scale | .03 | .02 | 0.08 | 0.03 |  |

Note: N= 1364; SE= standard error; CI= confidence interval.

### Mediational analysis

Two mediation analysis models were constructed: the first model included the sleep/circadian predictors self-reported sleep quality, circadian preference, and insomnia severity. In the next step, we added the covariates sex, age, marital status, education level, and employment status to create the second model.

#### Depression

Of the sleep/circadian predictors, only insomnia severity significantly mediated the association of ADHD symptom severity with depression severity (model 1: standardized beta = 0.05, bootstrapped *SE* = 0.008, bootstrapped 95% CI = [0.01, 0.06]). However, after adding the sociodemographic predictors, none of the sleep/circadian predictors (self-reported sleep quality, insomnia severity, and circadian preference) significantly mediated the association of ADHD symptom severity with depression severity(model 2: standardized beta = .049, bootstrapped *SE* = .009, bootstrapped 95% CI = [-0.03, 0.07]).

#### Quality of life

##### Satisfaction with Life Scale (SLS)

Of the sleep/circadian predictors, circadian preference and insomnia severity significantly mediated the association of ADHD symptom severity with SLS-related quality of life (model 1: standardized beta = -0.11, bootstrapped *SE* = 0.012, bootstrapped 95% CI = [-0.13, -0.09]). After adding the sociodemographic predictors, only insomnia severity significantly mediated the association of ADHD symptom severity with SLS-related quality of life (model 2: standardized beta = -0.10, bootstrapped *SE* = 0.011, bootstrapped 95% CI = [-0.13, -0.08]).

##### Cantril Ladder questionnaire (CL)

Of the sleep/circadian predictors, only insomnia severity significantly mediated the association of ADHD symptom severity with CL-related quality of life (model 1: standardized beta = .103, bootstrapped *SE* = .011, bootstrapped 95% CI = [0.08, 0.13]). Similarly, after adding the sociodemographic predictors, only insomnia severity significantly mediated the association of ADHD symptom severity with CL-related quality of life (model 2: standardized beta = .103, bootstrapped *SE 0*= .011, bootstrapped 95% CI = [0.08, 0.13]).

## DISCUSSION

We showed that ADHD symptom severity was associated with higher depression symptom severity, lower quality of life, insomnia severity, lower self-reported sleep quality and later circadian preference. Importantly, insomnia severity mediated the association of ADHD symptom severity with quality of life.

Approximately 30-35% of adults with ADHD experience at least one major depressive disorder episode throughout their lifespan, which is 15% higher than the risk observed in the general population ^23^. In a recent two-sample network Mendelian randomization analysis that aimed to identify mental disorders causally related to ADHD, the genetic likelihood of ADHD was associated with an increased risk of major depressive disorder^24^. Similarly, meta-analyses (ADHD: n = 550,748; no ADHD n = 14,546,814) yielded pooled odds ratios of 4.5 for major depressive disorder, suggesting that ADHD is highly comorbid with depression compared to non-ADHD^25^. Similarly, a combined longitudinal study (n = 8310) with two-sample Mendelian randomization analyses indicated childhood ADHD was associated with a higher risk of depression in adulthood, due to a causal effect of ADHD genetic likelihood on major depression^26^. Currently, there are no meta-analyses on Adult ADHD and quality of life. In a systematic review examining the association of adult ADHD and quality of life in six studies, ADHD diagnosis was consistently associated with an overall lower quality of life^27^. Similarly, in a multicenter study with young adults with ADHD and age- and sex-matched individuals with neurotypical development, ADHD was associated with lower quality of life^28^. Collectively, these findings suggest that ADHD is associated with an increased risk of depression and lower quality of life in adulthood. Here, we show similar associations of ADHD symptom severity with depression symptom severity and lower quality of life in the general population. Our findings thus help identify the importance of mental health and quality of life in individuals with ADHD traits, irrespective of a preexisting ADHD diagnosis. Individuals with ADHD are diagnosed with sleep disorders at a rate eight times higher than the general population^29^, manifesting in various ways such as delayed sleep onset, increased movement during sleep, daytime sleepiness, and shorter sleep duration^8^. A recent Swedish nationwide register-based study assessed sleep disorders, including insomnia, across five different age groups of ADHD and individuals with neurotypical development^29^. Accordingly, 7.5% of individuals with ADHD (n=145 490, 2.25% of the cohort), had a sleep disorder and 47.5% had been prescribed sleep medication. Irrespective of age, individuals with ADHD had an increased risk of having a sleep disorder diagnosis (OR range=6.4-16.1) and a sleep medication prescription, particularly for insomnia (OR range=12.0-129.4) compared with individuals with neurotypical development^29^. The circadian system may contribute to sleep disturbances, as up to a third of individuals with ADHD exhibit a late chronotype and delayed sleep-wake phase disorder^30^, with evidence for disrupted daily rhythms and circadian parameters ^31^. Here, we showed that ADHD symptom severity was associated with higher insomnia severity, lower self-reported sleep quality, and, to a lesser extent, with later circadian preference. Late circadian preference often occurs in individuals with ADHD who experience delayed sleep-wake phase disorder^13^. As the current study did not have information on delayed sleep-wake phase syndrome, this could explain the (relatively) weak association.

Importantly, we show that adult ADHD symptom severity and insomnia severity were significant predictors of lower quality of life and that insomnia severity mediated this association. However, we did not observe such findings for depression symptom severity. This difference could be due to the older age group in our study, which often reports lower depressive symptoms compared to younger age groups^32^ and/or the absence of clinically diagnosed depression compared to the studies above. To our knowledge, there are no previous reports of the association and mediation of sleep/circadian factors in depression and quality of life in ADHD. In one of the very few studies tangentially addressing this association in children and adolescents with ADHD (n=373, 10-19 years), self-reported sleep quality was lower in individuals with ADHD with mental disorders compared to those without^33^. Similarly, in a large cross-sectional study (n = 4618, aged 18-64), mental and physical functioning, work productivity, and healthcare use were adversely affected if ADHD diagnosis and insomnia symptoms co-occurred^34^. These findings combined suggest that insomnia severity may help explain the lower quality of life in individuals with ADHD traits.

Low sleep quality and self-reported insomnia occur in up to 60% of individuals with ADHD, which is 2.5-fold more than that for individuals with neurotypical development^35^. The etiology of SCRD in ADHD is complex and with limited understanding. Recent mechanistic insights suggest that insomnia risk genes are enriched in ADHD^36^. Disturbances in sleep and its timing are the rule, rather than the exception, across every diagnostic category of mental disorder^14^. SCRD is a risk factor for the subsequent development of mental disorders^37-39^, is one of the earliest signs of relapse^40^, and occurs both during and between acute episodes of illness^41^, where the severity of symptoms of sleep-circadian disruption and mental disorders co-vary. Given the critical link among ADHD symptom severity, insomnia severity, and mental disorders, future mechanistic studies including preclinical and human laboratory studies are needed to understand this complex interplay. Such understanding will be critical in the development of effective, scalable preventative and therapeutic interventions to improve the quality of life in individuals with ADHD traits. Moreover, it may help current clinical guidelines that do not link sleep/circadian complaints to quality of life in ADHD assessment.

Limitations of this study include the limited age range (primarily middle-aged adults) that limits translation to youth mental health and the cross-sectional design that does not allow testing for causal effects. In summary, our study shows that ADHD symptom severity was associated with lower quality of life, and this effect was mediated primarily by insomnia symptom severity. Future studies targeting insomnia complaints in individuals with ADHD traits may help mitigate mental health complaints and improve quality of life.

## Acknowledgements

The Netherlands Sleep Registry study was co-funded by the European Union (ERC-AdG Overnight, 101055383). Views and opinions expressed are however those of the authors only and do not necessarily reflect those of the European Union or the European Research Council. Neither the European Union nor the granting authority can be held responsible for them. The funder had no role in the design of the study; in the collection, analysis, or interpretation of data; in the writing of the manuscript, or in the decision to publish it.

## Author Contributions Statement

Conceptualization: E.V.S. Funding acquisition: E.V.S. Investigation: E.V.S. and S.L.C. Visualization: S.N., N.D., E.V.S., and S.L.C. Project administration: S.N., N.D., and S.L.C. Supervision: S.L.C. Data curation: S.N., N.D., E.V.S., and S.L.C. Formal analysis: S.N., N.D., and S.L.C. Validation: E.V.S., and S.L.C. Writing—review and editing: S.N., N.D., C.M.H., S.C., E.V.S., and S.L.C.

## Data Availability Statement

The data that support the findings of this study are available from the shared senior author and Principal Investigator, [E.V.S.], upon reasonable request.

